# Defining optimal anthropometric thresholds for sarcopenia screening in community-dwelling older Vietnamese adults

**DOI:** 10.1101/2025.08.03.25332901

**Authors:** Hien Thi Nguyen, Charuai Suwanbamrung, Apichai Wattanapisit, Warapone Satheannoppakao, Thang Nguyen, Tam Thai Thanh Tran, Khanh Hoang Pham, Dung Tam Nguyen Huynh, Cua Ngoc Le

## Abstract

Sarcopenia poses a growing health challenge for Vietnam’s aging population, yet effective screening remains limited by impractical standard diagnostics and a lack of validated, population-specific anthropometric thresholds. We conducted a cross-sectional study of 416 community-dwelling adults aged ≥60 years in Can Tho, Vietnam, to establish optimal cutoff values for body mass index (BMI), calf circumference (CC), arm circumference (AC), and waist circumference (WC) against the Asian Working Group for Sarcopenia 2019 (AWGS 2019) criteria. Receiver operating characteristic (ROC) analysis in R (pROC package) identified optimal cutoffs by maximizing the Youden index (sensitivity + specificity − 1): BMI ≤ 22.6 kg/m^2^ for men and ≤ 21.2 kg/m^2^ for women; CC ≤ 34.5 cm/32.0 cm; AC ≤ 26.0 cm/24.0 cm; and WC ≤ 85.0 cm/78.0 cm. Applying these Vietnam-specific thresholds improved the area under the curve from 0.75 to 0.77 for BMI, 0.61 to 0.70 for AC, 0.64 to 0.68 for CC, and 0.57 to 0.69 for WC, while specificity increased by 14–60% depending on the index. The Vietnam-specific BMI cutoff provided the best overall diagnostic accuracy; CC and AC were less effective in women, likely reflecting sex-specific differences in muscle mass and fat distribution, whereas WC achieved high specificity in women but lower overall accuracy, indicating potential utility of alternative measures such as the weight-adjusted waist index (WWI). These findings support the adoption of Vietnam-specific BMI thresholds as a simple, cost-effective primary screening tool for sarcopenia in older adults, with CC, AC, and WC as viable alternatives when BMI cannot be obtained. Implementing population-specific anthropometric cutoffs in national screening programs may facilitate earlier detection of sarcopenia, optimize resource allocation, and inform future research on combined indicator strategies.

## Introduction

Sarcopenia is a progressive geriatric syndrome characterized by the loss of skeletal muscle mass and function. Since 2016, it has been recognized as a distinct clinical condition under the International Classification of Diseases, 10th Revision, Clinical Modification (ICD-10-CM) code M62.84[1, 2]. In the context of a rapidly aging global population, sarcopenia has emerged as a significant public health issue due to its associations with increased risks of mortality, falls, hospitalization, disability, cognitive decline, and metabolic disorders[3–6]. People with sarcopenia have a fourfold increased risk of death and a threefold increased risk of functional decline compared to people without the condition. The prevalence of the disease varies widely, ranging from 5.2% to 62.7% in community settings and 25% to 73.7% in nursing homes [7].This condition places a significant burden on public health, affecting the physical and psychological well-being of older adults and placing significant pressure on the health care system. In the United States, medical costs associated with sarcopenia increased from $18.5 billion in 2000 to more than $40 billion in 2014, with an average cost of about $260 per person in 2019 [5, 8, 9].

Although dual-energy X-ray absorptiometry (DEXA) and bioelectrical impedance analysis (BIA) are considered gold standards for assessing muscle mass, their high cost and complex operations restrict their use widely. Anthropometric indices such as calf circumference (CC), and mid-upper arm circumference (AC) are simple, low-cost, and community-appropriate alternatives in primary care settings[10–12]. CC provides comparable accuracy, especially in women, and may not be affected by upper limb edema, making it an attractive single-measure screening [13, 14]. Other indices, such as BMI, waist circumference, or “Yubi-Wakka”, are simple but lack stability in obese or edematous individuals [15]. A systematic review found that simple, quick, non-invasive, and low-cost anthropometric measurements offered high diagnostic accuracy (pooled AUC: 0.84). Unlike questionnaires or physical performance tests, they were not influenced by dementia, subjective, or psychological factors. Their accuracy was slightly lower than multi-method tools like Ishii (pooled AUC: 0.89) [9].

Vietnam is facing a demographic shift, with the proportion of people aged ≥65 projected to double from 7% to 14% between 2020 and 2035, faster than most developed countries and ranking 3rd in ASEAN [16, 17]. Concurrently, changes in dietary habits, lifestyle, and extended life expectancy are contributing to increased risk of sarcopenia in the community. Existing studies report sarcopenia prevalence as high as 47% in community-dwelling elderly and up to 69% in institutionalized populations [18–20]. Despite this, the implementation of sarcopenia screening in Vietnam remains limited, particularly at the primary care level. A significant barrier to effective screening is the lack of validated, population-specific anthropometric thresholds. Relying on international thresholds derived from foreign populations, which exhibit notable variations in body composition, may result in substantial diagnostic bias. This can lead to overlooked cases if the threshold is excessively high or false positives if it is too low, thereby misallocating resources [21–23].

Therefore, locally verified standards should be identified for the sarcopenia screening program in Vietnam to ensure the effectiveness and logical resource allocation in public health programs. To close this research gap, the following research questions were posed for this study: (1) What are the optimal thresholds for BMI, CC, and AC for detecting sarcopenia in community-dwelling Vietnamese elderly? (2) How do these population-specific thresholds compare with the criteria of the Asian Working Group for Sarcopenia (AWGS) 2019 regarding diagnostic accuracy? In order to respond to these questions, this study sought to (i) identify specific CC, AC, WC, and BMI thresholds for sarcopenia screening among the Vietnamese elderly; and (ii) evaluate the diagnostic accuracy of those anthropometric indices according to the AWGS 2019 criteria. The study aims to provide practical, cost-effective, and ethnically appropriate screening tools for early detection and intervention. The results are expected to support public health strategies and national guidelines for aging populations in Vietnam.

## Materials and methods

### Study design and setting

This cross-sectional diagnostic accuracy study was conducted as part of the ViSarco Project, which aimed to improve sarcopenia screening among community-dwelling older adults in Vietnam. The research was conducted in Can Tho City, a central urban area in the Mekong Delta region. Data was collected at participants’ homes and the Can Tho University of Medicine and Pharmacy Hospital. The sarcopenia diagnosis was performed using the AWGS 2019 criteria, including DEXA, handgrip strength, and a 6-meter gait speed test. The study flowchart is presented in Figure 1.

**Figure 1.**
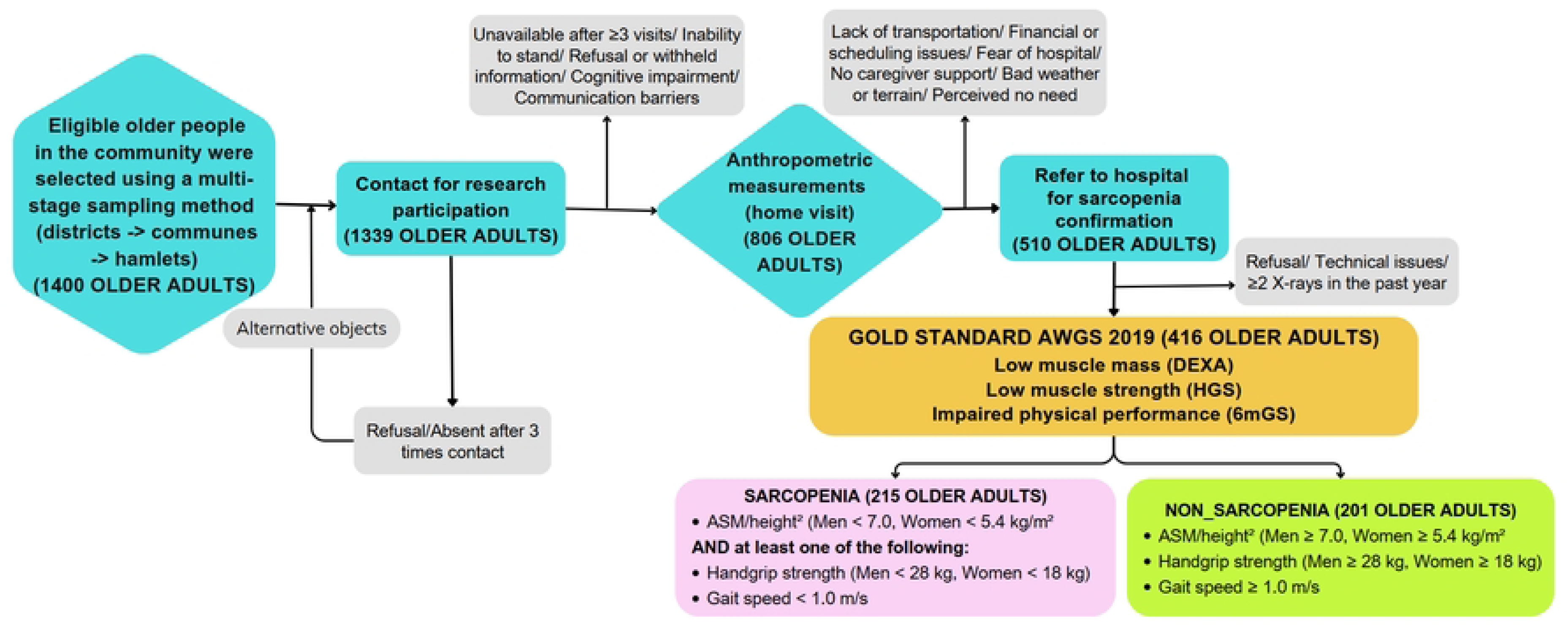
Research diagram.

### Study population and sampling

The process of sampling started on 2^nd^ September, 2025 and ended on 12^th^ October, 2025. We recruited participants using a multi-stage stratified random sampling technique. We purposively selected two districts in Can Tho, namely Ninh Kieu (urban) and Phong Dien (peri-urban), to capture demographic diversity. We randomly selected four communes or wards from each district, then randomly chose three hamlets within each. We randomly selected individuals aged 60 years or older from the household lists in each hamlet. To be eligible, participants had to reside in the community, walk independently, and provide informed consent. We excluded individuals who had experienced acute illness, hospitalization, or significant physical impairment within the past year. We contacted each eligible person up to three times; if they declined or were unavailable, we randomly selected a replacement from the same hamlet list. All procedures complied with the Declaration of Helsinki, and written informed consent was obtained from each participant. Before participation, all participants were informed about the study’s objectives, procedures, potential risks, and benefits. Written informed consent was obtained from all participants, ensuring their voluntary involvement and the right to withdraw at any time without any consequences.

### Sample size calculation

The required sample size was estimated using Buderer’s method[24], which is suitable for calculating sample size in diagnostic test accuracy studies based on expected sensitivity and specificity, desired confidence level, and acceptable margin of error [25].

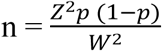

In this formula, n represents the required sample size; Z is the Z-score for 95% confidence level (1.96); p denotes the anticipated sensitivity or specificity and W is the desired precision (10% or 0.1).

Based on a previous Vietnamese study[26], the expected values were sensitivity at 66.7%, specificity at 67.1%, and sarcopenia prevalence at 66.1%. The minimum required sample size was 379 (129 for sensitivity, 250 for specificity). In practice, we enrolled 416 participants, 215 of whom had sarcopenia and 201 of whom did not, surpassing the minimum sample size.

### Measurements

Anthropometric measurements followed the standardized procedures of the Food and Nutrition Technical Assistance (FANTA) guidelines [27], We used a Seca 769 Digital Medical Scale to measure weight and height with participants barefoot and lightly clothed, then calculated BMI by dividing weight (kg) by height squared (m^2^). We measured AC and CC using a non-stretchable tape at the midpoint of the upper arm and the widest point of the calf, and measured WC at the midpoint between the iliac crest and lower ribs. We measured skeletal muscle mass by DEXA on a GE Lunar Prodigy scanner and classified lean mass as insufficient when the appendicular skeletal muscle mass index (ASM/height^2^) fell below 7.0 kg/m^2^ in men or 5.4 kg/m^2^ in women (AWGS 2019). To assess muscle strength, we seated participants with their shoulders adducted and neutrally rotated, elbows flexed at 90°, and wrists in a neutral position; we adjusted the Jamar hydraulic dynamometer handle to fit each hand, instructed them to squeeze as hard as possible for three seconds, allowed a 30-second rest between trials, and recorded the highest of three maximal efforts with the dominant hand [28]. We defined low strength as < 28 kg for men or < 18 kg for women. We then evaluated physical performance via two trials of a 6-meter usual-pace gait speed test, averaging the times, converting to m/s, and classifying poor performance as < 1.0 m/s. Finally, we diagnosed sarcopenia per AWGS 2019 by confirming low muscle mass plus either low muscle strength or poor physical performance [12].

### Data quality assurance

All field researchers were trained prior to data collection. Instruments were calibrated daily, and 10% of participants were randomly re-measured to assess inter-rater reliability. Data were double-entered and cross-verified for accuracy.

### Statistical analysis

This study used R software (version 4.3.1) to analyze the data. Using the pROC package, we assessed the diagnostic performance of anthropometric indices (BMI, CC, AC, WC) for sarcopenia via receiver operating characteristic (ROC) curves [29]. Briefly, we fit ROC models for each anthropometric index (BMI, CC, AC, WC) against sarcopenia status, extracted AUC, accuracy, sen, spec, PLR, and NLR with pROC::roc() and pROC::coords(), and then used cutpointr::cutpointr() specifying method = youden (which internally applies J = sen + spec – 1) to identify the threshold that maximized J [30]. AUC values were categorized into four accuracy levels for screening performance: low (0.50–0.70), acceptable (0.70–0.80), excellent (0.80–0.90), and outstanding (>0.90) [31]. Due to the absence of complete reference values in the Asian Working Group for Sarcopenia (AWGS) guidelines, we compared our results with benchmarks from populations of similar body composition: BMI (Japan) [32], AC (China) [31], CC (AWGS) [12], and WC (Amazon region) [33]. The full R script is presented in the supplementary (S1).

### Ethical approval

The Ethics Committee in Human Research of Walailak University approved the study (Approval No. WUEC-24-263-01, dated 24 July 2024).

## Results

The analysis of demographic and health characteristics of 416 participants stratified by sarcopenia status revealed stark contrasts between participants with and without sarcopenia. Sarcopenia was significantly more prevalent in males (64.4%) despite females comprising the majority of the sample (67.5%) (p < 0.001). It was also more common among older individuals and those with multiple chronic conditions (p < 0.05). Occupation showed a notable association, with higher rates observed in manual laborers and housekeepers.

Participants with sarcopenia had lower physical activity levels (p = 0.032) and significantly reduced BMI, waist, arm, and calf circumferences (all p < 0.001). No significant differences were detected in ethnicity or physical activity level categories (Table 1).

**Table 1.**
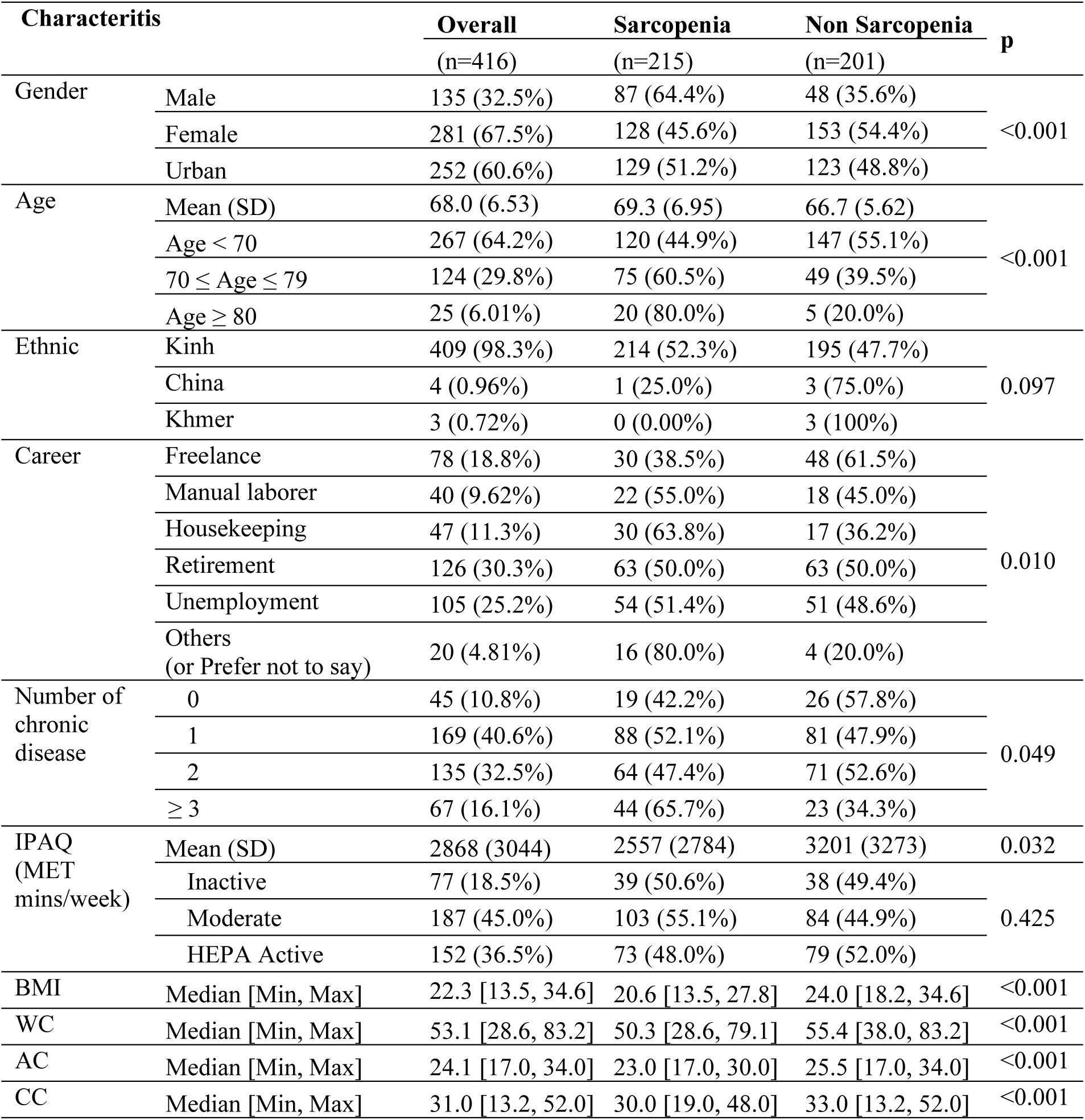
Demographic characteristics and some health indicators of study subjects according to sarcopenia status (n=416)

Table 2 summarizes the optimal cut-off values and diagnostic performance of four anthropometric indicators (BMI, CC, AC, and WC), identified using the Youden Index (J), which reflects the threshold with the highest combined sensitivity and specificity for each indicator. Across both genders, BMI demonstrated the strongest diagnostic performance (J = 0.51–0.58), followed by AC (J = 0.36–0.50), CC (J = 0.35–0.49), and WC showed the lowest utility (J = 0.27–0.36), especially in the combined-gender analysis. A complete list of cut-off values, along with their corresponding sensitivity and specificity rankings based on the Youden Index (J), is presented in supplementary (Table S1-4), and illustrated through sensitivity and specificity curves for anthropometric indices in supplementary (Fig S1).

**Table 2.**
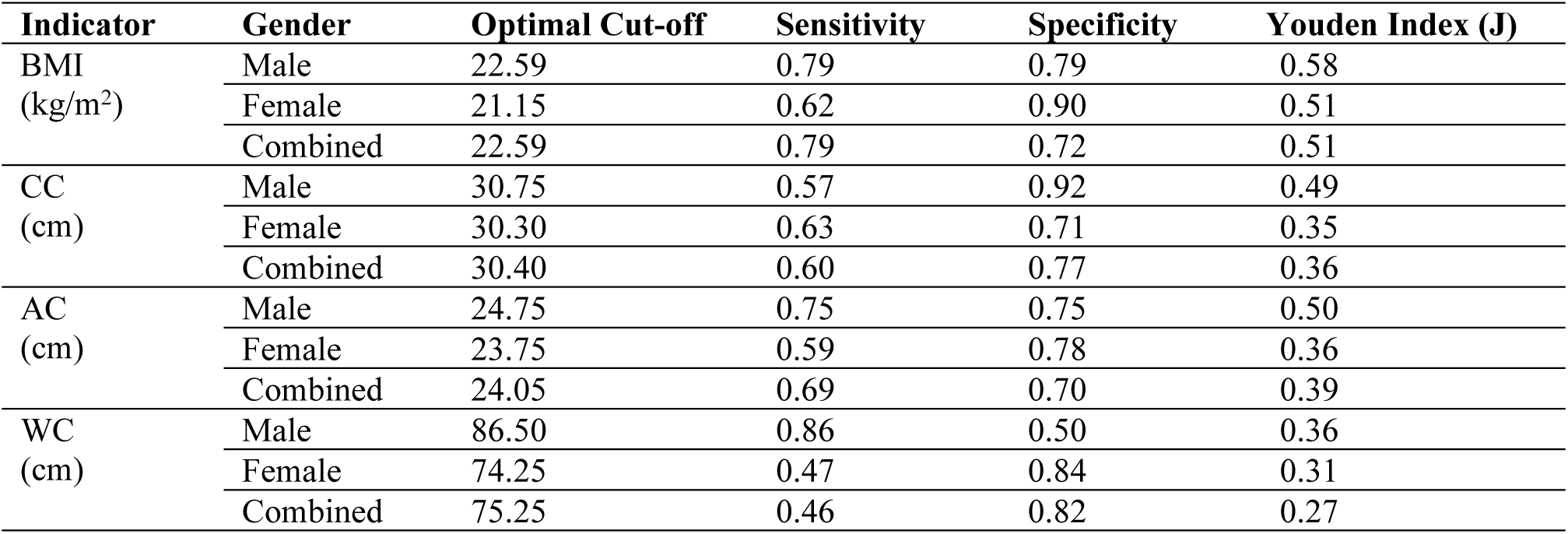
Optimal cut-off values and diagnostic performance of anthropometric indicators by gender.

When men and women were analyzed separately, every anthropometric measure achieved a higher Youden index than in the combined analysis. This outcome mirrors the physiological reality that women generally carry more body fat, possess less muscle mass, and distribute fat differently than men. In response, we implemented sex-specific cutoff values in order to enhance diagnostic accuracy. For practical application in clinical conditions, these values are rounded to the nearest appropriate unit. As a result, the proposed cut-off thresholds for the Vietnamese population are as follows: BMI *≤* 22.6 kg/m^2^ for men and 21.2 kg/m^2^ for women; CC *≤* 31 cm for men and 30 cm for women; AC *≤* 25 cm for men and 24 cm for women; and WC *≤* 86.5 cm for men and 74.5 cm for women.

When we applied the Vietnam-specific anthropometric cut-offs, all four indicators remained statistically significant predictors of sarcopenia after adjustment for age, sex, occupation, MET–activity time and number of chronic diseases. Using the reference (international) BMI threshold yielded an adjusted odds ratio (aOR) of 9.93 (95% CI: 6.22– 16.20; p < 0.001), whereas the Vietnam-specific BMI cut-off produced an even higher aOR of 13.73 (95% CI: 8.23–23.72; p < 0.001). This means that participants whose BMI fell below the Vietnam-specific threshold had roughly 13.7 times the odds of sarcopenia compared to those above the threshold.

Similarly, for calf circumference (CC) the aOR increased from 3.17 (95% CI: 2.04– 4.98) with the international cut-off to 4.38 (95% CI: 2.84–6.82) with the Vietnam-specific value, indicating a 4.4-fold higher odds of sarcopenia when CC is below 31 cm for men or 30 cm for women. Arm circumference (AC) and waist circumference (WC) showed parallel patterns: AC aOR rose from 3.14 to 4.78, and WC aOR from 3.29 to 5.84, all p < 0.001, underscoring that the locally derived thresholds better discriminate high-risk individuals in our population (Table 3).

**Table 3.** Comparison of adjusted odds ratios for sarcopenia risk based on reference and Vietnam-specific anthropometric cutoffs.

| Variable | Cutoff type | Predictor group<sup>#a</sup> | $\beta$ (SE) | aOR (95% CI)<sup>#b</sup> | p-value |
| --- | --- | --- | --- | --- | --- |
| BMI | Japan[32] | Low BMI | 2.295 (0.244) | 9.93(6.22-16.20) | <0.001 |
|  |  | Normal BMI |  | 1 (ref) |  |
|  | Vietnam cutoff | Low BMI | 2.620 (0.270) | 13.73 (8.23-23.72) | <0.001 |
|  |  | Normal BMI |  | 1 (ref) |  |
| CC | AWGS 2019[12] | Low CC | 1.152 (0.228) | 3.17 (2.04-4.98) | <0.001 |
|  |  | Normal CC |  | 1 (ref) |  |
|  | Vietnam cutoff | Low CC | 1.476 (0.223) | 4.38 (2.84-6.82) | <0.001 |
|  |  | Normal CC |  | 1 (ref) |  |
| AC | China[31] | Low AC | 1.143 (0.247) | 3.14 (1.95-5.13) | <0.001 |
|  |  | Normal AC |  | 1 (ref) |  |
|  | Vietnam cutoff | Low AC | 1.563 (0.222) | 4.78 (3.10-7.42) | <0.001 |
|  |  | Normal AC |  | 1 (ref) |  |
| WC | Amazon region[33] | Low WC | 1.191 (0.333) | 3.29 (1.75-6.50) | <0.001 |
|  |  | Normal WC |  | 1 (ref) |  |
|  | Vietnam cutoff | Low WC | 1.764 (0.250) | 5.84 (3.61-9.63) | <0.001 |
|  |  | Normal WC |  | 1 (ref) |  |

(#a)Reference cutoffs were based on previous population-specific studies (BMI: Japanese threshold $\leq 22.6$ kg/m<sup>2</sup> for both genders; CC: AWGS 2019 guideline $< 34$ cm for men, $< 33$ cm for women; AC: Chinese population values $< 25.9$ cm for men, $< 26.5$ cm for women; WC: Amazon region data $< 97$ cm for men, $< 86$ cm for women). Vietnam-specific cutoffs (derived from this study): BMI $\leq 22.6$ kg/m<sup>2</sup> (men), $\leq 21.2$ kg/m<sup>2</sup> (women); CC $\leq 31$ cm (men), $\leq 30.5$ cm (women); AC $\leq 25$ cm (men), $\leq 24$ cm (women); WC $\leq 86.5$ cm (men), $\leq 74.5$ cm (women).
(#b)Model adjusted for age, gender, career, MET, and number of chronic diseases.

Tailoring anthropometric thresholds to reflect Vietnamese population traits significantly improved sarcopenia diagnostic performance across all measurements (Table 4). For BMI, lowering the cutoff from the international reference of ≤ 22.6 kg/m^2^ for both sexes (Japan criteria) to ≤ 22.6 kg/m^2^ for men and ≤ 21.2 kg/m^2^ for women in Vietnam raised the AUC from 0.754 to 0.772, increased specificity by 14% (72→86 %), nearly doubled the positive likelihood ratio (2.79 → 4.77), and achieved an overall accuracy of 76.9 %.

**Table 4:**
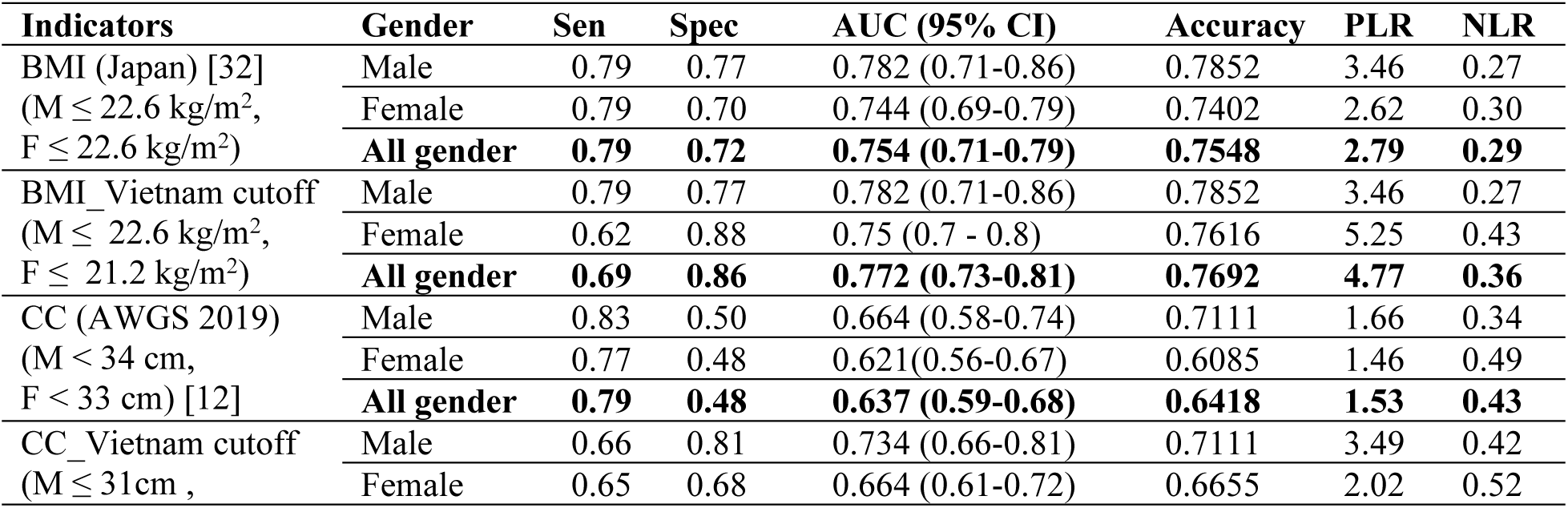
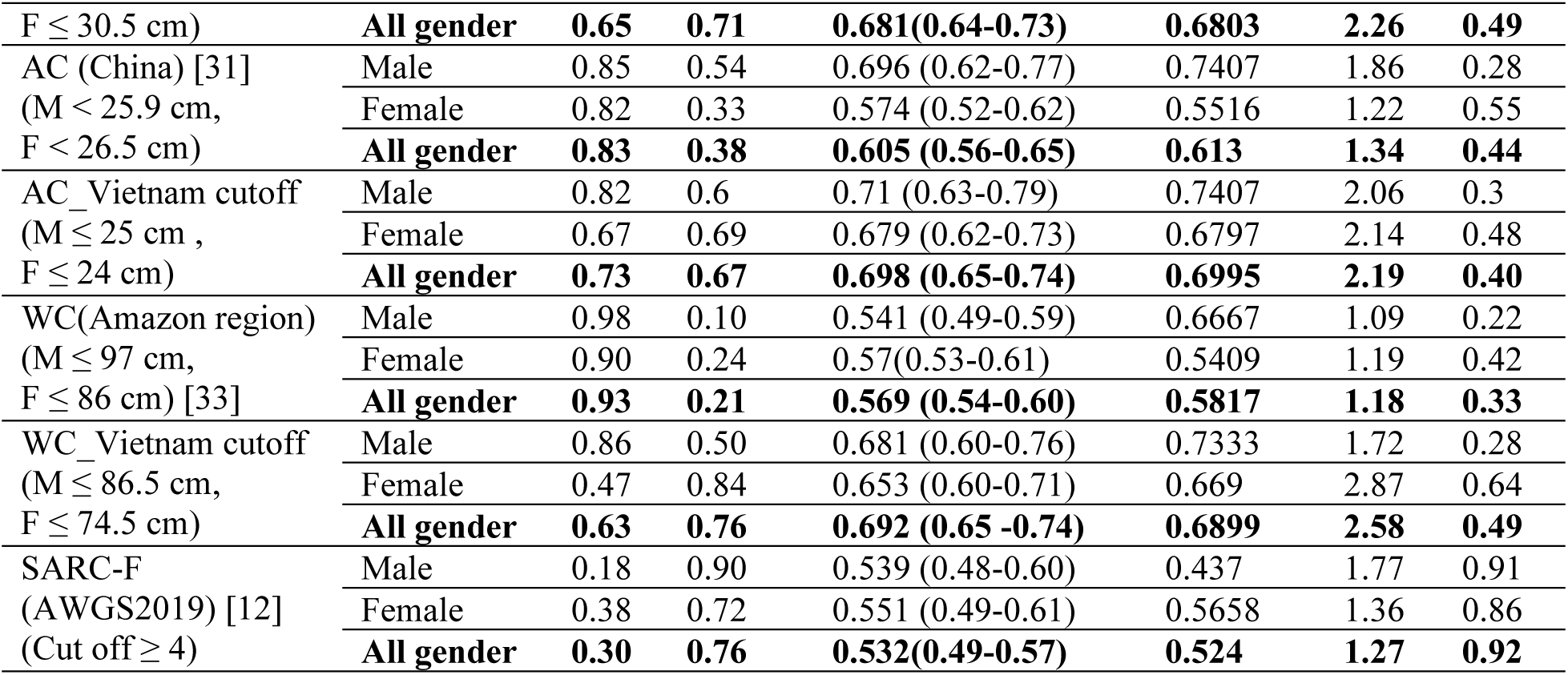
Diagnostic accuracy of an anthropometric index for sarcopenia in community-dwelling elders.

Similarly, CC cut-points were reduced from the AWGS 2019 standard (< 34 cm for men, < 33 cm for women) to ≤ 31 cm and ≤ 30.5 cm, respectively; this adjustment lifted the AUC to 0.681, improved specificity by 23% (48 →71 %), and yielded 68.0 % accuracy. AC thresholds fell from the Chinese standard (< 25.9 cm men, < 26.5 cm women) to ≤ 25 cm and ≤ 24 cm, boosting AUC to 0.698, specificity by 29% (38 → 67 %), and achieving 70.0 % accuracy. The most striking enhancement was seen in WC: lowering the Amazonian cutoffs (≤ 97 cm for men, ≤ 86 cm for women) to ≤ 86.5 cm and ≤ 74.5 cm increased AUC by 0.123 (0.569 → 0.692), surged specificity by 55% (21→76 %), and reached 69.0 % accuracy with female specificity leaping from 24 % to 84 %. Although these refined cut-points caused slight sensitivity declines, the marked gains in specificity, PLR, and overall accuracy underscore the critical value of population-specific criteria for effective sarcopenia screening in Vietnamese elders.

The ROC curve of anthropometric indices in the classification of sarcopenia in the elderly in Vietnam reflects the effectiveness of adjusting the threshold according to population characteristics. When applying the optimal threshold in Vietnam, BMI (AUC increased from 0.754 to 0.772), CC (AUC from 0.637 to 0.681), AC (AUC from 0.605 to 0.698), and WC (AUC from 0.57 to 0.692) all showed significant improvements in disease discrimination (Fig. 2).

**Figure 2.**
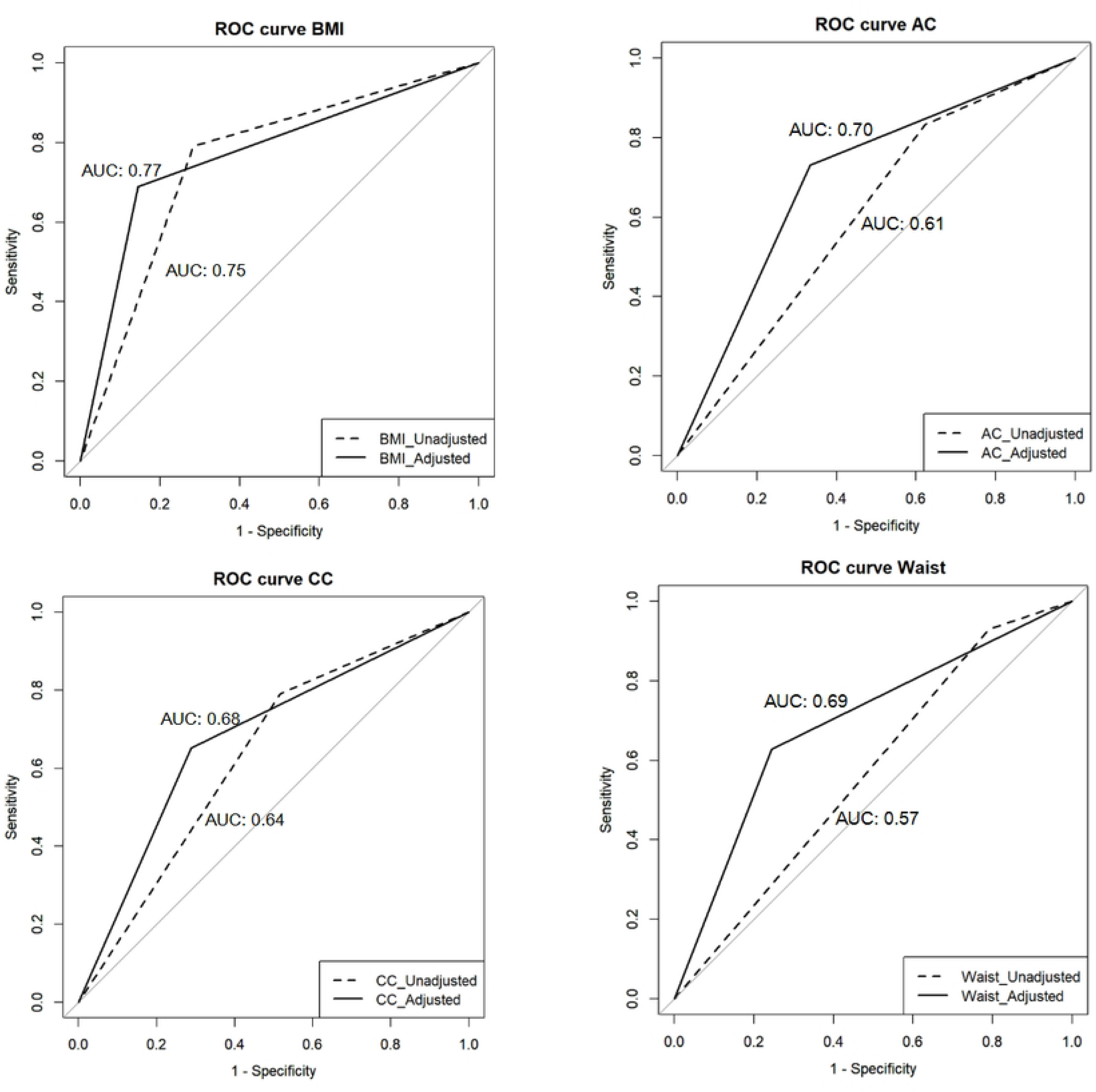
ROC curve of anthropometric indices for classifying sarcopenia in elderly Vietnamese.

## Discussion

### Value of Ethnically Adapted Anthropometric Thresholds

The study showed that all anthropometric indices were capable of discriminating sarcopenia in the elderly in the community in Vietnam (AUC > 0.5), with superior accuracy compared to the SARC-F, a validated and widely used tool. After modifying the cutoffs for the Vietnamese population, the diagnostic performance of sarcopenia was greatly improved for all anthropometric indices, emphasizing the significance of developing suitable criteria for every ethnic group. However, the classification level was still limited to low as acceptable. In particular, the Vietnamese-specific BMI threshold showed the highest screening performance for sarcopenia in both sexes (AUC = 0.772 (0.73–0.81)), the best among the indices (close to the good discrimination level)., and a high positive likelihood ratio (PLR = 4.77), a high PLR significantly increasing clinical confidence in positive screening results.The diagnostic performance improvements achieved by ethnically adapted anthropometric thresholds highlight the importance of population-specific standards for sarcopenia screening. This result aligns with findings from a study in Thailand (AUC: 0.74)[34] but is lower than those reported in China (AUC >0.8)[35]. Despite all being Asian populations, our BMI cutoff is lower than both Thailand and China (Thailand: <24; China: <25 kg/m^2^ for both sexes). These variations reflect population-specific differences in anthropometric traits and body fat distribution. The lower cutoff reflects the unique nutritional and physical profiles of Asian populations. Prior research has shown that, at the same BMI, Asians tend to have higher levels of body fat and visceral fat compared to Caucasians, an attribute often described as the ’thin-fat’ phenotype [36].

Meanwhile, CC and AC, two indices that are often prioritized and have high accuracy in Asia due to their association with lean body mass, have lower performance in Vietnam, CC and AC give acceptable results in men (AUC 0.734 and 0.71, respectively) but low in women (AUC 0.664 and 0.679), reflecting gender differences in performance. This reflects the heterogeneity in nutritional status, physiological characteristics, and fat mass distribution even among Asian countries, requiring adjustment of cutoff points or assessment methods to suit each local context. The Vietnamese people’s cutoff index in our study was lower than that of the Chinese and AWGS, which is consistent with China’s assessment that this value may not be suitable for Asian older adults and needs to be adjusted according to the region [31].

The difference in diagnostic performance of sarcopenia based on anthropometric indices between the two sexes stems from physiological differences in the proportion and distribution of muscle and fat between men and women, in addition to the fact that postmenopausal women have a decrease in estrogen, which increases visceral fat accumulation and reduces muscle mass, making anthropometric indices less sensitive in detecting sarcopenia in women [37].

The WC cutoff for the Vietnamese population demonstrated relatively low discriminatory power, with AUC values ranging from 0.653 to 0.692 and accuracy between 66.9% and 73.3%. Despite limited overall performance, specificity in women was notably high at 84%. These findings are lower than those reported in a Brazilian study, where WC showed stronger predictive ability for sarcopenia (AUC > 0.70) [38]. However, they are comparable to results from a Japanese study, which reported an AUC of approximately 0.515. This suggests that WC may not be an optimal proxy for muscle mass, likely due to the confounding effect of visceral fat in the abdominal region [39]. Notably, recent data from the U.S. National Health and Nutrition Examination Survey (NHANES) introduced the weight-adjusted waist index (WWI)—calculated as WC divided by the square root of body weight— as a more sensitive indicator of sarcopenia (OR 14.55 in men; OR 2.86 in women) [40]. However, this index has not been widely applied in Vietnam, warranting further investigation in future studies. The optimal WC threshold identified for Vietnamese women (≤74.5 cm) was notably lower than the Asian IDF standard by approximately 10 cm, reflecting the specific "thin but atrophied" phenotype common in Vietnam, where muscle atrophy may occur without significant central obesity. Such substantial differences reinforce the need for separate thresholds to prevent misclassification, analogous to ethnically tailored criteria already widely recognized in metabolic syndrome screening [41].

Besides that, despite being widely used and validated in Vietnam, the SARC-F questionnaire had very low sensitivity (18–38%), which resulted in poor diagnostic performance (AUC < 0.6). In comparison, earlier research conducted in the hospital found higher AUCs (0.71–0.72) [26], possibly because five questions of SARC-F focus only on muscle function and are more suitable for detecting severe muscle atrophy commonly seen in hospitalized patients. Therefore, although functional or symptom-based tools such as the SARC-F are simple and easy to use, they appear to be inadequate when used alone for screening in the community setting in Vietnam.

### Implications for Public Health Policy and Clinical Practice

Implementing simple, accurate, and population-specific screening methods, such as the Vietnamese BMI thresholds, would significantly enhance early detection of sarcopenia at primary care and community levels [23].Early identification facilitates timely lifestyle interventions, such as targeted physical activity and nutritional support, which can slow muscle loss and reduce risks associated with sarcopenia, including falls, functional decline, hospitalization, and reduced quality of life[12, 23].Using Vietnamese-specific BMI cutoffs (≤22.6 kg/m^2^ for men and ≤21.2 kg/m^2^ for women) as initial screening tools optimizes resource utilization, especially in settings without advanced body composition assessment equipment. A high PLR makes clinicians more confident in positive screening outcomes, guiding the appropriate selection of further diagnostic evaluations.

Furthermore, the use of cost-effective anthropometric screening tools can optimize healthcare resource allocation by reserving more expensive methods (DEXA, BIA) for those identified as high risk during initial screening [23]. Avoiding inaccurate international thresholds helps prevent unnecessary follow-ups due to low specificity or missed opportunities from low sensitivity.

These findings provide robust scientific support for developing national clinical guidelines and public health strategies in Vietnam, incorporating specific thresholds tailored to the elderly Vietnamese population. To fully realize these benefits, concerted efforts must include educating healthcare providers about sarcopenia, integrating these specific anthropometric thresholds into clinical protocols, and developing culturally appropriate health education materials.

This study is limited in the geographic area and demographic specificity. The sample was drawn exclusively from Can Tho City in the Mekong Delta region, with limited representation of Vietnam’s broader ethnic and regional diversity. As such, the applicability of the identified anthropometric cut-off points (BMI, CC, AC, WC) may not generalize to other provinces, particularly mountainous or northern regions, or ethnic minority groups with different body composition profiles and lifestyle factors. While the cut-offs demonstrated strong screening performance within the Can Tho population, particularly for BMI, they require further validation in larger, multi-regional, and multi-ethnic cohorts. Additionally, the reduced sensitivity of CC and AC in women suggests that sex-specific or combined-indicator approaches (e.g., BMI + CC) should be explored to enhance screening sensitivity. Finally, external validation and longitudinal studies are needed to assess the predictive accuracy of these thresholds over time.

## Conclusion

Overall, the Vietnam-specific BMI thresholds (≤22.6 kg/m^2^ for men and ≤21.2 kg/m^2^ for women) have demonstrated strong potential as practical initial screening tools for age-related loss of sarcopenia in older adults, offering high diagnostic accuracy (AUC: 0.75– 0.77), favorable positive likelihood ratios (PLR), and practical utility in clinical practice.

Other anthropometric indicators, including CC, AC, and WC, also showed improved diagnostic value after local threshold adjustments (CC: ≤31/30.5 cm; AC: ≤25/24 cm; WC: ≤86.5/74.5 cm), with particular effectiveness observed in men (e.g., CC and AC). However, these measures alone may be insufficient, particularly for women, due to reduced discriminatory power. These results underline the need for more study to optimize multi- parameter techniques and evaluate their function in promoting healthy aging, and they justify the inclusion of locally adjusted cutoff values in national screening guidelines.

## Data Availability

All relevant data are within the manuscript and its Supporting Information files

## Acknowledgments

This study was supported by the Graduate Scholarship from Walailak University, Thailand (Contract No. 07/2023). We also thank Can Tho University of Medicine and Pharmacy and all collaborators in Vietnam for their support in data collection.

## Source of funding

This research work was financially supported by Walailak University Graduate Research Fund, Contract No. CGS-RF-2024/18.

## Conflict of interest

The authors declare that there are no conflicts of interest.

## Ethical approval

This study is part of the ViSarco Project, a multi-phase research initiative aimed at improving sarcopenia screening strategies in Vietnam. This institution-based original study received ethical approval from the Ethics Committee in Human Research of Walailak University on 24 July 2024, under approval number WUEC-24-263-01.

## Author contributions

**Conceptualization:** Hien Thi Nguyen, Charuai Suwanbamrung, Cua Ngoc Le

**Methodology:** Hien Thi Nguyen, Thang Nguyen, Cua Ngoc Le

**Data collection:** Khanh Hoang Pham, Tam Thai Thanh Tran, Dung Tam Nguyen Huynh

**Data curation:** Thang Nguyen, Dung Tam Nguyen Huynh

**Formal analysis:** Hien Thi Nguyen, Apichai Wattanapisit, Warapone Satheannoppakao

**Visualization:** Apichai Wattanapisit, Warapone Satheannoppakao

**Project administration:** Hien Thi Nguyen, Cua Ngoc Le

**Funding acquisition:** Charuai Suwanbamrung, Cua Ngoc Le

**Writing – original draft:** Hien Thi Nguyen, Khanh Hoang Pham, Tam Thai Thanh Tran

**Writing – review and editing:** Hien Thi Nguyen, Cua Ngoc Le

## Supporting information

**S1 file: Detailed R script for Youden Index and diagnostic accuracy** (DOCX)

**S1– S4 Table. Cut-off Values Derived from the Youden Index and Corresponding Sensitivity and Specificity for AC, BMI, CC, WC**(DOCX)

**S1 Fig.**
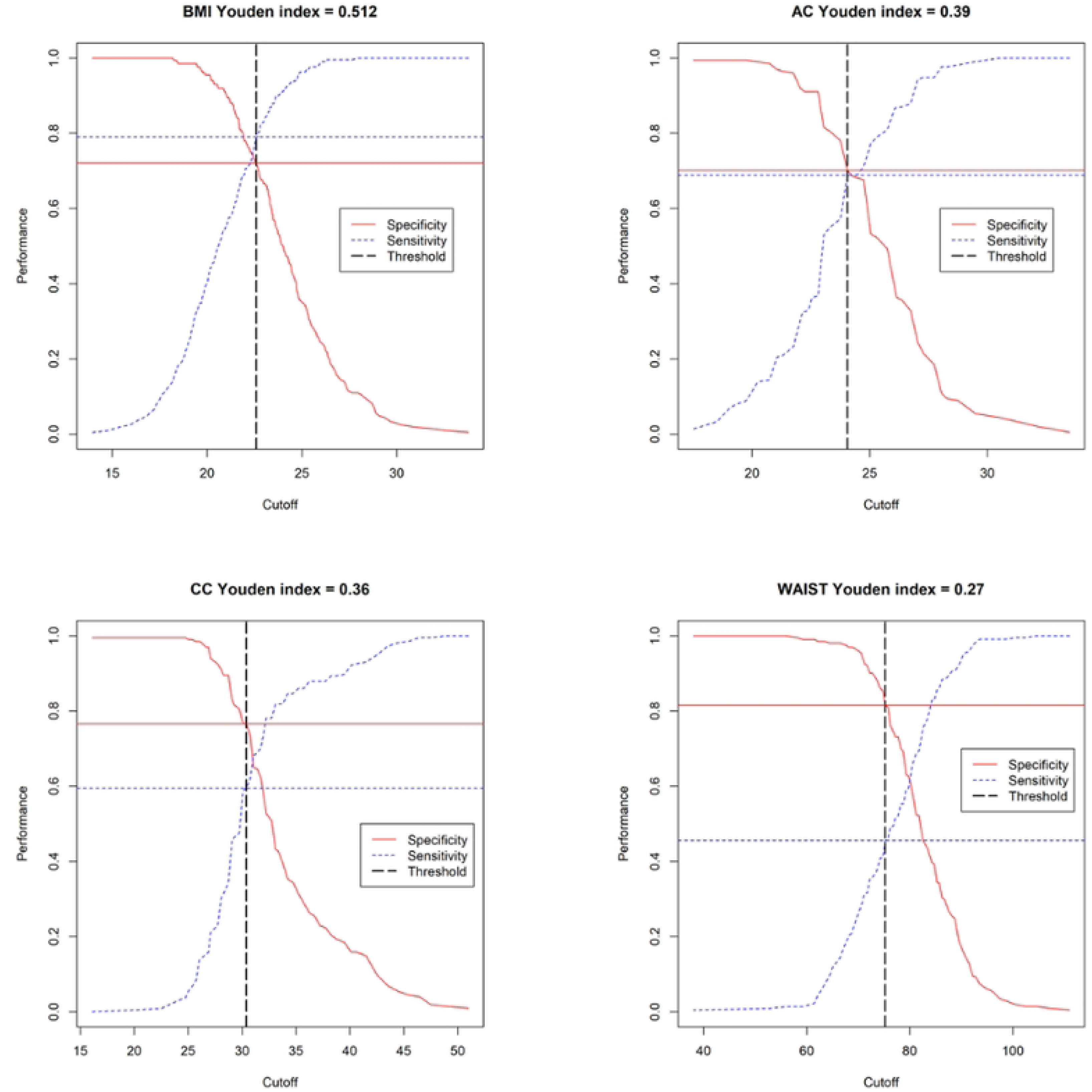
Sensitivity and Specificity Curves by Cut-off Values for Anthropometric Indices (Youden Index Method)

## Notes

### Competing Interest Statement

The authors have declared that no competing interests exist

### Funding Statement

Yes

